# Middle Temporal Gyrus Neurite Microstructure and Scam Susceptibility in Older Adults Without Dementia

**DOI:** 10.64898/2026.11.11.26367846

**Authors:** Melanie Leguizamon, Belinda Y. Zhang, Rachel M. Custer, Cassidy P. Molinare, Emma Oyen, Daisy T. Noriega-Makarskyy, Jenna Axelrod, Aaron C. Lim, Camdyn Wu, Maanya Agarwal, Lauren E. Salminen, Jeiran Choupan, Laura Mosqueda, S. Duke Han

## Abstract

**Background:** Financial exploitation is a prevalent public health concern among older adults, and emerging evidence links vulnerability to gray matter macrostructure and white matter microstructure, including within brain regions and networks implicated in early Alzheimer’s disease. Although diffusion tensor imaging is sensitive to age-related variation in brain microstructure, its scalar metrics cannot distinguish among the tissue properties contributing to observed differences. Neurite orientation dispersion and density imaging (NODDI) models distinct tissue compartments, yielding estimates of neurite density, dendritic complexity, and extracellular free water.

**Methods:** This study examined associations between scam susceptibility and NODDI metrics using ultra-high-field 7-Tesla imaging in 116 older adults without dementia. Neurite density index (NDI), orientation dispersion index (ODI), and free-water fraction were quantified within seven a priori regions of interest linked to early Alzheimer’s disease. Multivariate regressions evaluated associations between scam susceptibility and NODDI metrics, adjusting for age, sex, education, and global cognition, with multiple comparison correction within each metric. For significant regions, post-hoc hierarchical regression models tested whether NODDI metrics explained variance beyond regional cortical thickness, total gray matter, and total intracranial volume.

**Results:** Greater scam susceptibility was associated with higher NDI (*β*=0.30, *p_FDR_*=.025) and lower ODI (*β*=-0.28, *p_FDR_*=.044) in the middle temporal gyrus (MTG). No other regions survived correction. Additionally, MTG NDI and ODI explained variance beyond macrostructure measures.

**Conclusions:** These findings identify NODDI metrics as novel microstructural correlates of scam susceptibility beyond macrostructure, supporting investigation of their relevance to financial vulnerability in older age.

## 1. Introduction

Financial exploitation is a serious public health concern among older adults (Peterson et al., 2014), with prevalence rates doubling between 2008 and 2022 (Leguizamon et al., 2026). As the older adult population grows (Shreya et al., 2025) and technology use continues to evolve (Zhang et al., 2023), there are increasing opportunities and methods for older adults to be targeted by scams. Beyond the immediate financial toll, exploitation is associated with long-lasting negative effects on mental, emotional, and physical health (Nguyen et al., 2021). Given these consequences, identifying factors associated with financial exploitation vulnerability has become an important research endeavor.

While significant cognitive decline (Boyle et al., 2012) and mild cognitive impairment (Han, Boyle, James, et al., 2016) have been associated with scam susceptibility, many older adults may show financial vulnerability without any apparent cognitive deficits (Fenton et al., 2022; Hanoch et al., 2025; Lachs & Han, 2015). Moreover, greater scam susceptibility, or an increased tendency to fall for deceptive schemes, has been associated with an increased risk of future incident Alzheimer’s disease (AD; Boyle et al., 2025). Together, these findings suggest that scam susceptibility represents a behavioral correlate of subtle neural changes that may emerge alongside, or even precede, overt cognitive impairment or dementia.

Consistent with this possibility, previous neuroimaging research has identified macrostructural correlates of scam susceptibility in older adults without dementia. In models adjusted for cognitive ability, greater scam susceptibility was associated with lower gray matter density in the hippocampus, parahippocampal gyri, and middle temporal gyri (Han, Boyle, Yu, et al., 2016), as well as lower cortical thickness in entorhinal, fusiform, and middle temporal cortices (Lamar et al., 2024). These regions are vulnerable to early AD-related neuropathology (Buckner et al., 2008; Dickerson et al., 2009), aligning with prior evidence linking financial vulnerability to subsequent AD risk. However, this previous work has primarily examined macrostructural measures using voxel-based and volume-based morphometry techniques, which yield limited information about the organization of tissue at the microstructural level.

Diffusion-weighted imaging offers complementary information about brain microstructure by measuring the displacement of water molecules within tissue (Alexander et al., 2007). Conventional metrics include fractional anisotropy and mean diffusivity, which reflect the directional coherence and magnitude of water diffusion, respectively, and are most commonly assessed in white matter (Pierpaoli et al., 1996). In models adjusted for cognitive ability, greater scam susceptibility was associated with lower fractional anisotropy in right temporoparietal white matter pathways (Lamar et al., 2020), suggesting that microstructural white matter abnormalities may contribute to vulnerability to scams. Neurite Orientation Dispersion and Density Imaging (NODDI; Zhang et al., 2012) extends conventional diffusion imaging by modeling diffusion signals attributed to the intra-neurite, extra-neurite, and cerebrospinal fluid environments of the tissue. The model provides estimates of intracellular neurite fraction (NDI), variability in neurite orientations (ODI), and isotropically diffusing water (FISO). Although NODDI is often applied to white matter, its compartment-based indices may be particularly informative in gray matter, where diffusion is less directionally organized and conventional metrics may have limited biological specificity (Greenman & Bennett, 2025). Moreover, examining gray matter NODDI may help characterize local differences in neurite density, organization, and extracellular free water within cortical regions supporting the cognitive and socioemotional processes involved in detecting scams. Thus, whereas prior white matter findings implicate alterations in the connections between regions, gray matter NODDI may provide complementary and more sensitive information about the neurite density, organization, and extracellular free water within the implicated regions themselves.

The present study examined associations between scam susceptibility and NODDI-derived gray matter measures within seven a priori cortical and subcortical regions of interest (ROIs). These regions were selected based on their vulnerability to early AD (Buckner et al., 2008; Dickerson et al., 2009; Marra et al., 2021; Vonk et al., 2020), and included the hippocampus, entorhinal cortex, medial orbitofrontal cortex, parahippocampal gyrus, posterior cingulate cortex, precuneus, and middle temporal gyrus (MTG). As prior gray matter studies have generally associated aging and neurodegeneration with lower cortical NDI and ODI and higher FISO (Li et al., 2025; Xie et al., 2026; X. Yu et al., 2024), we hypothesized that greater scam susceptibility would be associated with lower NDI and ODI and higher FISO across our seven a priori ROIs. NDI and ODI were designated as primary metrics of interest. FISO was evaluated as a secondary metric because it is less specific to the underlying tissue process and is particularly sensitive to cerebrospinal fluid (CSF) partial volume effects in gray matter. For regions showing significant associations, exploratory post-hoc analyses examined whether NODDI metrics explained variance in scam susceptibility beyond regional and global measures of brain macrostructure.

## 2. Methods

### 2.1. Participants

Participants included adults aged 50 years and older recruited through online advertisements, community outreach events, and local senior centers across the greater Los Angeles area, as well as through National Center on Elder Abuse educational events and the Alzheimer’s Prevention Registry (Langbaum et al., 2020). Inclusion criteria were English fluency and being free of dementia (no self-reported diagnosis of dementia, a priori cutoff score of ≥ 21 on the modified 5-minute telephone-screened Montreal Cognitive Assessment; Wong et al., 2015). Exclusion criteria were any self-reported neurological or psychiatric disease, history of alcohol use disorder within the past five years, or history of other substance use disorder. All participants provided informed consent per the local Institutional Review Board and consistent with the Declaration of Helsinki and its amendments.

A total of 142 participants underwent brain imaging and completed neuropsychological assessment, including the susceptibility to scams scale. Four participants were excluded due to missing diffusion-weighted imaging data, eight because of FreeSurfer reconstruction errors, 14 due to poor image acquisition or quality (e.g., excessive head motion or inadequate slice coverage), and two because of diffusion processing errors. The final analytic sample comprised 116 participants.

### 2.2. Susceptibility to scams scale

Scam susceptibility was evaluated using the self-report Susceptibility to Scams (James et al., 2014) questionnaire, in which participants rated their agreement to five statements about phone habits and general scam knowledge. Responses were recorded on a Likert scale ranging from 1 = strongly agree to 7 = strongly disagree. Total scores were calculated as an average of responses across items, with certain items reverse scored, and ranged from 1 to 7. Higher scores indicated greater scam susceptibility. The intraclass coefficient for the measure has been reported as 0.63 (James et al., 2017), and higher scores on this scale have been associated with poorer semantic memory and financial and health literacy (Yu et al., 2021).

### 2.3. Imaging acquisition

Whole brain magnetic resonance imaging (MRI) data were acquired on a 7-Tesla Siemens Magnetom scanner equipped with an 8-channel parallel transmit coil. A 3D T_1_-weighted magnetization-prepared-rapid-gradient-echo (MPRAGE) sequence was acquired with repetition time (TR) = 2200 ms, echo time (TE) = 2.95 ms, flip angle = 7°, voxel size = 0.7 × 0.7 × 0.7 mm, 240 sagittal slices. DWI data were acquired using a 2D spin-echo echo-planar imaging sequence with CMRR multiband acceleration [TR = 6000 ms, TE = 66 ms, voxel size = 1.3 x 1.3 x 1.3 mm, field of view = 210 mm, partial Fourier = 6/8] and consisted of multi-shell b-values of approximately 700, 1350, and 2000 s/mm^2^ across 67 diffusion directions, along with 11 interspersed b0 images (78 volumes total). Anterior–posterior and posterior– anterior encoding directions were acquired and used for susceptibility distortion correction. Parallel imaging was implemented using GRAPPA (acceleration factor = 3) and simultaneous multi-slice acquisition (multiband factor = 2).

### 2.4. Diffusion processing

DWI data were preprocessed using FSL (version 6.0.7) in accordance with the ENIGMA-DTI guidelines (https://github.com/ENIGMA-git/ENIGMA-DTI-Preprocessing-Guidelines). Preprocessing included denoising, Gibbs ringing correction, susceptibility-induced distortion correction using TOPUP, and eddy current and head motion correction using EDDY_OPENMP. Preprocessed data were subsequently analyzed using the Quantitative Imaging Toolkit (QIT; (Cabeen et al., 2018). NODDI microstructural metrics were estimated using a multi-compartment spherical-mean approach implemented in QIT. The use of ultra-high-field 7-Tesla imaging further provided increased spatial resolution and signal-to-noise ratio, thereby reducing partial-volume contamination in our a priori selected ROIs (Platt et al., 2021).

### 2.5. NODDI parameters and regions of interest

NODDI parameters included NDI, ODI, and FISO. To reduce the influence of CSF partial volume in gray matter ROIs, tissue-weighted regional means were calculated for NDI and ODI by weighting each voxel according to its estimated tissue fraction, defined as 1 – FISO (Parker et al., 2021). Gray matter ROIs were defined using the FreeSurfer Desikan-Killiany cortical parcellation (Desikan et al., 2006) and subcortical segmentation (Fischl, 2012). Regional NODDI metrics were calculated for seven a priori selected ROIs: hippocampus, entorhinal cortex, medial orbitofrontal cortex, parahippocampal gyrus, posterior cingulate cortex, precuneus, and MTG. These regions were selected based on their established involvement in functions relevant to scam susceptibility and vulnerability to early neuropathological changes in AD (Buckner et al., 2008; Dickerson et al., 2009). Left- and right-hemisphere measures were averaged to create bilateral values for each ROI.

### 2.6. Macrostructural measures

T_1_-weighted images were processed using the high-resolution reconstruction pipeline implemented in FreeSurfer version 7.2.0 (Zaretskaya et al., 2018). Cortical thickness was estimated according to the Desikan-Killiany parcellation. Left- and right-hemisphere values were averaged to obtain a bilateral measure. FreeSurfer reconstructions were visually inspected, and segmentation errors were manually corrected when necessary. Corrected outputs underwent a final visual inspection, and scans that did not meet quality control criteria were excluded. Total gray matter volume and estimated total intracranial volume (eTIV) were also derived from FreeSurfer outputs.

### 2.7. Statistical analyses

Standard descriptive statistics were calculated for all demographic and behavioral variables of interest. Separate multiple linear regression models examined the association between scam susceptibility and each NODDI measure (NDI, ODI, and FISO) within each ROI. All continuous variables were standardized as z-scores before analysis. Models were adjusted for age, sex, years of education, and global cognition measured using the MMSE. We corrected for multiple comparisons across the seven a priori ROIs separately for each NODDI metric using the false discovery rate (FDR) Benjamini-Hochberg procedure (Benjamini & Hochberg, 1995).

### 2.9. Post-hoc analyses

Exploratory post-hoc analyses were conducted to further characterize NODDI findings that survived FDR correction in the primary analyses. Hierarchical regression models evaluated whether the identified regional NDI and ODI measures explained variance in scam susceptibility beyond conventional measures of brain macrostructure. In regional macrostructure models, age, sex, years of education, MMSE score, and bilateral ROI cortical thickness were entered in Block 1, with NDI or ODI entered in Block 2. In global macrostructure analyses, age, sex, years of education, MMSE score, total gray matter volume, and eTIV were entered in Block 1, followed by either NDI or ODI in Block 2. Nested models were compared using partial *F*-tests to evaluate the change in explained variance (Δ*R*²).

## 3. Results

### 3.1. Participants

Participant characteristics are summarized in Table 1. The sample consisted of predominantly female (77.59%), White (64.66%), and highly educated (16.37 ± 2.08 years) individuals free of dementia, with an average MMSE score of 28.41 ± 1.54 and an average scam susceptibility score of 1.91 ± 0.80.

**Table 1.**
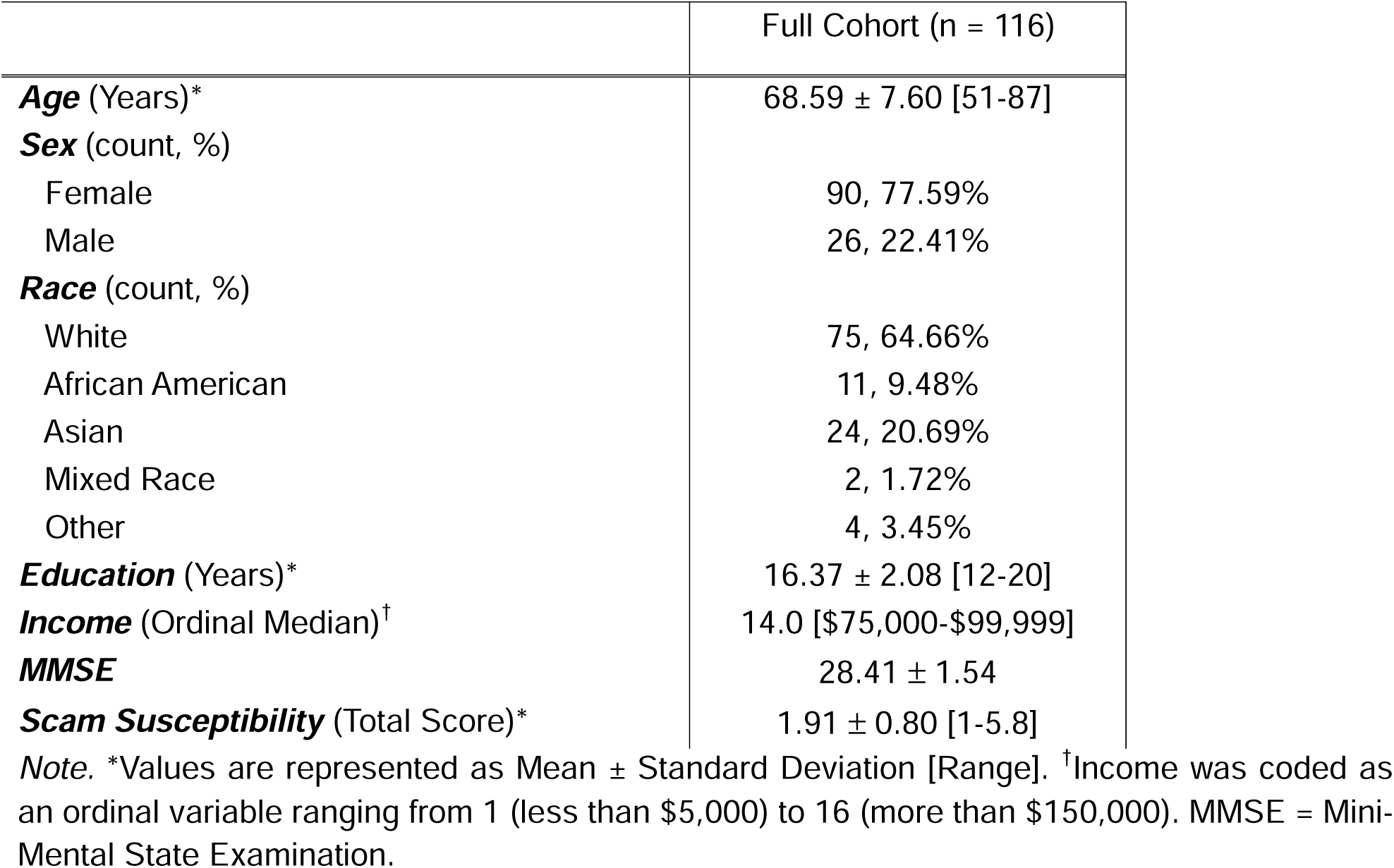
Sample characteristics.

|  | Full Cohort (n = 116) |
|---|---|
| ***Age*** (Years)<sup>\*</sup> | 68.59 ± 7.60 [51-87] |
| ***Sex*** (count, %) |  |
| Female | 90, 77.59% |
| Male | 26, 22.41% |
| ***Race*** (count, %) |  |
| White | 75, 64.66% |
| African American | 11, 9.48% |
| Asian | 24, 20.69% |
| Mixed Race | 2, 1.72% |
| Other | 4, 3.45% |
| ***Education*** (Years)<sup>\*</sup> | 16.37 ± 2.08 [12-20] |
| ***Income*** (Ordinal Median)<sup>†</sup> | 14.0 [\$75,000-\$99,999] |
| ***MMSE*** | 28.41 ± 1.54 |
| ***Scam Susceptibility*** (Total Score)<sup>\*</sup> | 1.91 ± 0.80 [1-5.8] |

*Note.* <sup>\*</sup>Values are represented as Mean ± Standard Deviation [Range]. <sup>†</sup>Income was coded as an ordinal variable ranging from 1 (less than \$5,000) to 16 (more than \$150,000). MMSE = Mini-Mental State Examination.

### 3.2 Associations between NODDI metrics and scam susceptibility

After correcting for multiple comparisons, greater scam susceptibility was associated with higher NDI (*β* = 0.30, 95% CI [0.10, 0.49], *p* = .004, *p_FDR_* = .025) and lower ODI (*β* = -0.28, 95% CI [-0.48, -0.08], *p* = .006, *p_FDR_*= .044) in the MTG (Figure 1; Table 2). Higher ODI in the precuneus was also nominally associated with greater scam susceptibility (*p* = .042) but did not remain significant after FDR correction. NODDI metrics were not significantly associated with scam susceptibility for the other a priori ROIs.

**Figure 1.**
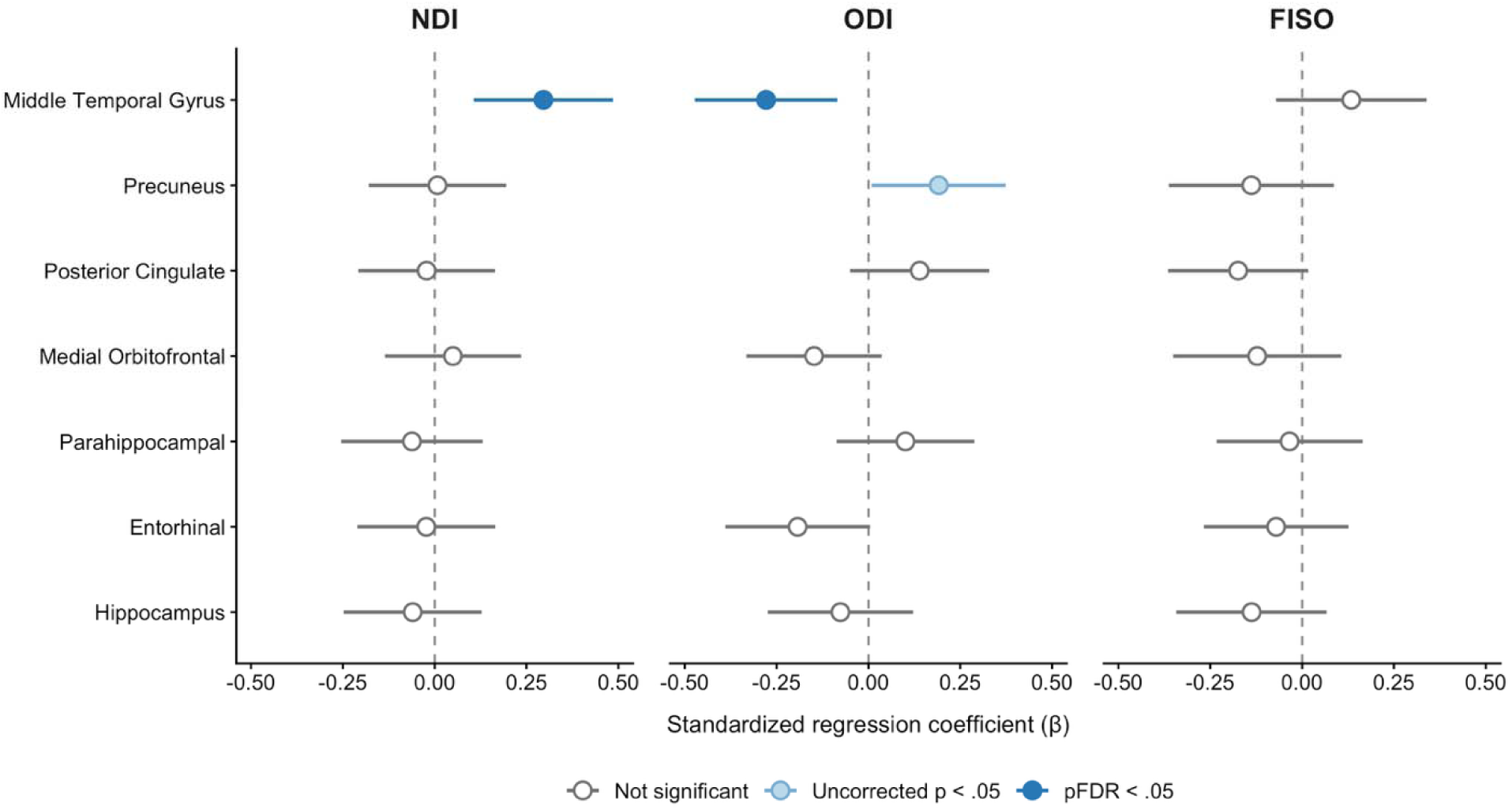
Regional associations between NODDI metrics and scam susceptibility. *Note.* Points represent standardized regression coefficients and horizontal lines represent 95% confidence intervals. Models were adjusted for age, sex, years of education, and global cognition (MMSE). Dark blue points indicate associations that remained significant following FDR correction performed separately across the seven regions within each NODDI metric. Light blue points indicate nominal associations with uncorrected *p* < .05 that did not survive multiple comparison correction, and open gray points indicate associations with uncorrected *p* > .05. NDI = neurite density index; ODI = orientation dispersion index; FISO = isotropic volume fraction; FDR = false discovery rate.

**Table 2.** Adjusted associations between regional NODDI metrics and scam susceptibility.

|  | **NDI** |  |  | **ODI** |  |  | **FISO** |  |  |
|---|---|---|---|---|---|---|---|---|---|
| Region | *β* | *p* | *p<sub>FDR</sub>* | *β* | *p* | *p<sub>FDR</sub>* | *β* | *p* | *p<sub>FDR</sub>* |
| Middle Temporal Gyrus | **0.23** | **.003** | **.020** | **-0.28** | **.006** | **.041** | 0.13 | .204 | .403 |
| Precuneus | 0.01 | .937 | .937 | 0.19 | .042 | .134 | -0.14 | .230 | .403 |
| Posterior Cingulate | -0.02 | .817 | .937 | 0.14 | .151 | .212 | -0.17 | .076 | .403 |
| Medial Orbitofrontal | 0.05 | .601 | .937 | -0.15 | .118 | .207 | -0.12 | .298 | .417 |
| Parahippocampal | -0.06 | .530 | .937 | 0.10 | .295 | .344 | -0.03 | .735 | .735 |
| Entorhinal | -0.02 | .811 | .937 | -0.19 | .057 | .134 | -0.07 | .483 | .563 |
| Hippocampus | -0.06 | .534 | .937 | -0.08 | .451 | .451 | -0.14 | .190 | .403 |

*Note.* Models adjusted for age, sex, education, and global cognition (MMSE). FDR correction was performed separately across the eight regions within each NODDI metric. Boldface indicates associations that remained significant after FDR correction (*p<sub>FDR</sub>* < .05). *β* = standardized regression coefficient; NDI = neurite density index; ODI = orientation dispersion index; FISO = isotropic volume fraction; FDR = false discovery rate.

### 3.3 Post-hoc analyses

Separate hierarchical regression models were conducted to determine whether NDI and ODI in the MTG explained variance in scam susceptibility beyond conventional gray matter macrostructural metrics previously examined in relation to scam susceptibility (Han, Boyle, Yu, et al., 2016; Lamar et al., 2024). In these post-hoc analyses, NDI and ODI remained associated with scam susceptibility after adjustment for bilateral MTG cortical thickness. Specifically, adding NDI explained an additional 7.6% of variance, *F*(1, 109) = 9.43, *p* = .003, and adding ODI explained an additional 8.0% of variance, *F*(1, 109) = 10.02, *p* = .002 (Table 3). Results were also robust to adjustment for total gray matter volume and eTIV, with NDI and ODI explaining an additional 5.6% and 5.3% of variance, respectively (NDI: *F*(1, 108) = 6.91, *p* = .010; ODI: *F*(1, 108) = 6.46, *p* = .012; Table 3).

**Table 3.** Contribution of middle temporal gyrus microstructural metrics to scam susceptibility beyond macrostructural brain measures.

| Macrostructural comparison | Metric Added | *β* | Δ*R*<sup>2</sup> | *F* | *p* |
|---|---|---|---|---|---|
| Total GM volume + eTIV | NDI | .261 | .056 | 6.91 | **.010** |
| MTG cortical thickness | NDI | .295 | .076 | 9.43 | **.003** |
| Total GM volume + eTIV | ODI | -.258 | .053 | 6.46 | **.012** |
| MTG cortical thickness | ODI | -.312 | .080 | 10.02 | **.002** |

*Note.* Block 1 in all models included age, sex, years of education, and global cognition (MMSE). Global volume models additionally included total gray matter volume and total intracranial volume, while regional models instead included mean middle temporal gyrus cortical thickness. NDI or ODI were adding in Block 2. Continuous variables were standardized. The reported *F* and *p* values correspond to the nested-model comparison testing the addition of the NODDI metric. *β* = standardized regression coefficient; MTG = middle temporal gyrus; NDI = neurite density index; ODI = orientation dispersion index; GM = gray matter; eTIV = estimated total intracranial volume. Significant p-values are bolded.

## 4. Discussion

To our knowledge, this study is the first to investigate NODDI-derived gray matter microstructural metrics in relation to scam susceptibility among older adults without dementia. Greater scam susceptibility was associated with higher NDI and lower ODI in the MTG after adjustment for age, sex, education, and global cognition and multiple comparison correction. No other regions showed significant associations with scam susceptibility after FDR correction. In post-hoc analyses, NDI and ODI in the MTG explained variance in scam susceptibility beyond macrostructural measures. Together, these findings suggest that scam susceptibility may reflect MTG microstructural alterations that are not detectable through conventional macrostructural measures.

The association between lower ODI in the MTG and greater scam susceptibility may be consistent with reduced complexity or dispersion in gray matter neurite organization (Gozdas et al., 2021; Shi et al., 2025) and aligns with prior evidence of lower ODI with advancing age and neurodegeneration (Nir et al., 2024; X. Yu et al., 2024). Contrary to our hypothesis, greater scam susceptibility was also associated with higher MTG NDI. Although NDI is commonly interpreted as an estimate of neurite density, higher cortical NDI may also arise from cellular swelling associated with toxic metabolite accumulation, or from reductions in extracellular space (Sacco et al., 2020; Xie et al., 2026). Considered jointly, higher NDI and lower ODI may reflect gray matter tissue with a greater apparent intra-neurite fraction but less branching or variability in neurite orientation. Such a profile could result from selective loss or simplification of neuritic processes, while the higher NDI may arise from neuroinflammation, reduced extracellular space, or other tissue changes that increase the apparent intra-neurite signal fraction (Dowell et al., 2019).

Nonlinear or biphasic structural trajectories observed in the earliest stages of neurodegenerative disease further support the possibility that increases in a structural metric may precede subsequent decline (Montal et al., 2021). Because the MTG supports semantic processing and retrieval relevant to evaluating the plausibility of scams (Davey et al., 2016), reduced microstructural complexity in this region could interfere with the efficient recognition and interpretation of deceptive cues. Greater scam susceptibility may therefore be associated with an altered MTG microstructural architecture characterized by relatively high apparent intra-neurite fraction but reduced organizational complexity. However, this interpretation remains preliminary, as the biological substrate of this NODDI profile cannot be established from the present data alone and requires validation using longitudinal imaging, complementary biomarkers, and histological studies.

Exploratory post-hoc analyses further demonstrated that MTG NDI and ODI explained variance in scam susceptibility beyond regional cortical thickness, total gray matter volume, and intracranial volume. These findings extend prior structural neuroimaging research implicating temporal regions in financial vulnerability. Previous studies have linked greater scam susceptibility to lower gray matter volume in the MTG (Han, Boyle, Yu, et al., 2016), lower cortical thickness in AD-vulnerable entorhinal, fusiform, and middle temporal regions (Lamar et al., 2024), and lower white matter fractional anisotropy in middle temporal regions (Lamar et al., 2020). The present findings suggest that NODDI captures aspects of MTG microstructure related to scam susceptibility that are not fully accounted for by conventional structural measures. However, the biological characteristics underlying these associations remain unclear.

Several limitations should be acknowledged. First, the sample consisted of predominantly White, highly educated, and female participants, limiting generalizability to more diverse populations. Validating these findings in larger, more demographically diverse samples is an important direction for future work. Second, the susceptibility to scams scale is a five-item self-report measure that may not fully capture future risk of financial exploitation and is subject to recall bias. Longitudinal studies tracking changes in NODDI-derived microstructure among individuals who go on to experience a scam are needed to establish whether these associations reflect risk prospectively. Third, given the cross-sectional study design, causal relationships between microstructure and scam susceptibility cannot be established. Fourth, we did not have access to amyloid, tau, or other biomarker data to confirm whether the observed microstructural differences reflect underlying Alzheimer’s disease-related pathology or normal variation across aging. Future studies incorporating biomarker data would help clarify whether these microstructural profiles are associated with preclinical neurodegenerative processes.

## 5. Conclusion

In conclusion, this is the first study to evaluate NODDI-derived gray matter microstructural metrics in relation to scam susceptibility in older adults without dementia using ultra-high-field 7-Tesla imaging. Higher NDI and lower ODI in the MTG were associated with greater susceptibility to scams and explained variance beyond regional cortical thickness, total gray matter volume, and total intracranial volume. Together, these findings identify MTG NODDI metrics as novel microstructural correlates of scam susceptibility and support longitudinal investigation into whether they are sensitive to changes that precede macrostructural atrophy or cognitive decline.

## Data Availability

All data produced are available online at https://doi.org/10.5281/zenodo.24142660.

## Ethics statement

This study was approved by the local Institutional Review Board and consistent with the Declaration of Helsinki.

## Data and code availability statement

The investigators from the current study will fully comply with any and all NIH policies to share data and code in order to support the aims of greater scientific inquiry, diversity of empirical approach, training of new researchers, and testing of new or alternative hypotheses. De-identified data and code will be made available to qualified researchers through a transparent and fair process after contacting the PI and submitting a formal request for data. All requests for data, database, and code submissions will be documented for record-keeping purposes.

## CRediT authorship contribution statement

**Melanie Leguizamon:** Conceptualization, Methodology, Formal analysis, Writing – original draft, Writing – review and editing, Visualization; **Rachel M. Custer:** Methodology, Software, Writing – review and editing; **Belinda Y. Zhang:** Conceptualization, Writing – review and editing; **Cassidy P. Molinare:** Writing – review and editing; **Emma Oyen:** Writing – review and editing; **Daisy T. Noriega-Makarskyy:** Writing – review and editing; **Jenna Axelrod:** Writing – review and editing; **Aaron C. Lim:** Conceptualization, Writing – review and editing; **Camdyn Wu:** Data curation, Writing – review and editing; **Maanya Agarwal:** Data curation, Writing – review and editing; **Lauren E. Salminen:** Conceptualization, Supervision, Writing – review and editing; **Jeiran Choupan:** Methodology, Software, Writing – review and editing**; Laura Mosqueda:** Writing – review and editing; **S. Duke Han:** Conceptualization, Supervision, Funding acquisition, Writing – review and editing.

## Acknowledgements

We thank participants from the greater Los Angeles community for taking part in our study.

